# Enhancing Ischemic Stroke Management: Leveraging Machine Learning Models for Predicting Patient Recovery After Alteplase Treatment

**DOI:** 10.1101/2024.11.05.24316803

**Authors:** Babak Khorsand, Atena Vaghf, Vahide Salimi, Maryam Zand, Seyed Abdolreza Ghoreishi

## Abstract

**Aim:** Ischemic stroke remains a leading global cause of morbidity and mortality, emphasizing the need for timely treatment strategies. This study aimed to develop a machine learning model to predict clinical outcomes in ischemic stroke patients undergoing Alteplase therapy, thereby supporting more personalized care.

**Methods:** Data from 457 ischemic stroke patients were analyzed, including 50 demographic, clinical, laboratory, and imaging variables. Five machine learning algorithms—k-nearest neighbors (KNN), support vector machines (SVM), Naïve Bayes (NB), decision trees (DT), and random forest (RF)—were evaluated for predictive accuracy. The primary evaluation metrics were sensitivity and F-measure, with an additional feature importance analysis to identify high-impact predictors.

**Results:** The Random Forest model showed the highest predictive reliability, outperforming other algorithms in sensitivity and F-measure. Furthermore, by using only the top-ranked features identified from the feature importance analysis, the model maintained comparable performance, suggesting a streamlined yet effective predictive approach.

**Conclusion:** Our findings highlight the potential of machine learning in optimizing ischemic stroke treatment outcomes. Random Forest, in particular, proved effective as a decision-support tool, offering clinicians valuable insights for more tailored treatment approaches. This model’s use in clinical settings could significantly enhance patient outcomes by informing better treatment decisions.

## 1. Introduction

Ischemic stroke stands as one of the most devastating neurological emergencies and a life-threatening medical condition, often leading to severe long-term disability or even death [1]. According to estimates, 1 in 4 adults over the age of 25 experience an ischemic stroke in their lifetime, underscoring the widespread impact of this condition [2]. This condition arises from the sudden death of brain tissue due to a dramatic reduction in blood flow and oxygen supply to the brain, often caused by the formation of clots and occlusion of cerebral arteries [3].

Risk factors for ischemic stroke can be classified into non-modifiable and modifiable categories. Non-modifiable risk factors include age, sex, and ethnicity, whereas modifiable risk factors encompass hypertension, smoking, poor dietary habits, and physical inactivity. The clinical presentation of ischemic stroke often includes sudden onset of paralysis, numbness, or weakness on one side of the body, difficulties with speech, visual disturbances, loss of balance, dizziness, severe headaches, and vomiting [4, 5]. Prompt recognition and early intervention are vital, as timely treatment can significantly improve functional recovery and overall outcomes following a stroke event.

Tissue plasminogen activator (tPA), particularly Alteplase, is marked as the gold standard for ischemic stroke therapy. It has been shown to effectively reverse or mitigate the effects of ischemic stroke when administered within 4.5 hours of symptom onset. However, the efficacy and safety of tPA have been controversial, particularly regarding its use in patients with later presentation than the recommended time window [6-8].

In recent years, computational approaches have significantly aid in simulating brain function, assessing stroke-induced damage, and modelling drug interactions [9-11]. Machine Learning (ML) has emerged as a powerful tool in the realm of medical decision-making, offering a data-driven approach that enhances clinical practices. ML algorithms excel in pattern recognition, which reduces human bias and improves the accuracy of predictions [12-17]. Their applications in the field of ischemic stroke are expanding, including in the diagnosis, personalized treatment decision-making, and prediction of complications and patient outcomes following treatment [18]. Early and accurate predictions of both the disease and treatment outcomes are crucial, as they empower clinicians to implement optimal care strategies, ultimately reducing the risk of adverse outcomes, and enhancing the quality of life for stroke survivors [19].

Given the significance of monitoring patients after receiving Alteplase and the uncertainties surrounding the safety and efficacy of Alteplase in cases of delayed presentation, we aimed to develop a robust machine□learning model to predict the potential effects of Alteplase therapy in ischemic stroke patients. This model is supposed to assist clinicians in making informed decisions regarding patient management.

## 2. Material and Method

### 2.1. Study Design and Patients

This retrospective study analysed the clinical data of 457 ischemic stroke patients (253 females and 204 males) admitted to Valiasr Hospital of Zanjan, Iran in 2021. The patients, aged between 29 and 99 years (mean age: 69.49 ± 10 years), received Alteplase therapy during hospitalization. The study aimed to develop a machine learning (ML) model to predict the efficacy of Alteplase in ischemic stroke patients based on various clinical and demographic attributes.

### 2.2. Data Collection

A total of 50 attributes (features) were collected for each patient, including demographic information, medical history, treatment data, and stroke characteristics. These features were recorded at admission and during hospitalization, covering the following categories:

#### 2.2.1. Demographic and clinical data

- Gender, Age, Weight
- Time of Stroke Onset
- Smoking Status
- History of Pharmaceutical Treatment (Yes/No):
  ▪ Anti-platelet medication
  ▪ Oral anticoagulants
  ▪ Heparin
  ▪ Oral anti-diabetic medication
  ▪ Insulin Therapy
  ▪ Anti-hypertensive medication
  ▪ Statins
- Medical History (Yes/No)
  ▪ Diabetes
  ▪ Hyperlipidaemia
  ▪ Previous stroke or transient ischemic attack (TIA)
  ▪ Atrial fibrillation
  ▪ Type of atrial fibrillation (transient/permanent)
  ▪ Congestive heart failure
  ▪ Vascular disease

#### 2.2.2. Laboratory and Imaging Data

- Blood glucose concentration at admission and 24 hours post-admission
- Activated partial thromboplastin time (APTT)
- Serum creatinine levels
- Neuroimaging data
  ▪ CT (imaging-computed tomography)
  ▪ MRI (magnetic resonance imaging)
- Systolic and diastolic blood pressure at admission and 24 hours post-admission

#### 2.2.3. Medication Administered During Hospitalization

- Aspirin or/and Clopidogrel
- Pantoprazole or/and Carvedilol or/and Nitrocontin

#### 2.2.4. Stroke Characteristics

Stroke characteristics were evaluated based on the National Institutes of Health Stroke Scale (NIHSS). NIHSS evaluates stroke severity across multiple domains:

- NIH1A: Level of Consciousness (LOC)
- NIH1B: LOC questions (birthday and age recall)
- NIH1C: LOC commands (eye and hand movements) (nih1A_baseline)
- NIH2: Best gaze (horizontal eye movements) (nih2)
- NIH3: Visual function (finger counting or visual threat)
- NIH4: Facial Palsy
- Motor function
  ▪ NIIH5A: right arm movement
  ▪ NIH5B: left arm movement
  ▪ NIH6A: right leg movement
  ▪ NIH6B: left leg movement
- NIH7: Limb ataxia (coordination and tremor evaluation)
- NIH8: Sensory function (sensory loss evaluation)
- NIH9: Best language (object identification, repetition, and speech production)
- NIH10: Dysarthria (speech intelligibility and/or speech naturalness)
- NIH11: Extinction and inattention (inferred from other tests)

NIHSS sub-scores and total scores were calculated to quantify the overall stroke severity.

### 2.3. Machine Learning Algorithms

To predict clinical outcomes after Alteplase therapy, five popular supervised ML algorithms were employed: k-nearest Neighbors (KNN), Support Vector Machine (SVM), Naïve Bayes (NB), Decision Trees (DT), and Random Forest (RF).

#### 2.3.1. k-Nearest Neighbors (kNN)

The kNN is a simple yet effective classification algorithm that classifies a sample based on the majority class of its ***k*** nearest neighbor, determined using the Euclidean distance, which measures the similarity between samples. The value of ***k*** determines the number of neighbors considered for classification [20].

#### 2.3.2. Support Vector Machine (SVM)

The SVM method is recognized as a benchmark classifier in artificial intelligence. This method constructs a decision boundary, or hyperplane, between the two closest training samples to separate the samples into the two classes by maximum marginal distance and minimum classification errors. SVM assumes that the number of dimensions exceeds the number of samples to reduce classification errors [21].

#### 2.3.3. Naïve Bayes (NB)

Naïve Bayes is a probabilistic learning model based on Bayes’ theorem. It operates under the assumption that the features are independent. This classifier uses the maximum probability principle to classify input data into specific classes [22].

#### 2.3.4. Decision Tree (DT)

DT is a well-established ML technique used for classification and prediction. A decision tree is visualized as a tree-like graph comprising a root node as well as several internal nodes and leaf nodes. To make predictions, the algorithm selects the optimal feature at each step, navigating the tree until it reaches a leaf node that indicates the class for a sample [23].

#### 2.3.5. Random Forest (RF)

Random Forest improves prediction accuracy by combining several decision trees. It generates smaller trees using random subsets of features and employs a majority voting strategy across all the trees to predict the class label of a sample [24].

### 2.4. Evaluation Metrics

The performance of the ML models was evaluated using several classification metrics derived from the confusion matrix: sensitivity, specificity, accuracy, and the F1-score [25].

A confusion matrix categorizes prediction into four outcomes: true positive (TP, number of instances correctly predicted as positive), true negative (TN, number of instances correctly predicted as negative), false positive (FP, number of instances incorrectly predicted as positive), and false-negative (FN, number of instances incorrectly predicted as negative).

#### 2.4.1. Sensitivity (Recall)

Sensitivity, also known as true positive rate, quantifies the proportion of actual positive samples that are correctly identified by the model. This metric is calculated using the formula [26]:

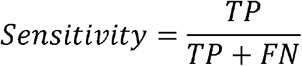

#### 2.4.2. Specificity

Specificity, or the true negative rate, evaluates the proportion of negative samples that are correctly classified [27]. This metric is particularly important where false positives can lead to unnecessary interventions. Specify is calculated as:

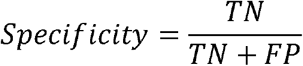

#### 2.4.3. F1-score

The F1-score, also known as f-measure, balances Precision (precision=TP/TP+FP) and Recall, providing a harmonic mean of two. The F1-score is calculated as [28]:

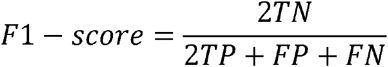

### 2.5. Feature Selection

To enhance the predictive capabilities of our models and quantify the contribution of input features to predict clinical outcomes, a backward elimination strategy was adopted. In this approach, an initial full model is trained with all variables (features). The variables are eliminated individually from the full model until only those that meaningfully influence the outcome remain [29, 30]. Five popular algorithms including kNN, SVM, NB, DT, and RF were employed to construct the models. After omitting each feature, they were assigned a score based on their contribution to predicting patient outcomes. Next, important features were identified and the models were retrained and tested using the top features. Finally, the evaluation metrics were calculated for the models, providing insight into model performance and the accuracy of the predictions.

## 3. Results

### 3.1. Initial Model Training

We started by training five predictive models, KNN, SVM, NB, DT, and RF, using fifty features. The performance metrics of models are summarized in Table 1 and illustrated in Figure 1. Both SVM and RF had a sensitivity of 0.97, indicating their highest performance in detecting positive instances. In contrast, NB showed a notably lowest sensitivity of 0.7 suggesting the limitation of this classifier for capturing true positives compared to other models. However, NB performed the best than its counterparts in terms of specificity, having a score of 0.93. This suggests that NB was the most effective model at correctly identifying true negatives, thereby minimizing false positive predictions. On the other hand, KNN showed a challenge in reducing false positives since it had the lowest specificity score.

**Table 1:** Predictive performance of KNN, SVM, NB, DT, and RF.

| Predictive model | Model performance |  |  |  |
| --- | --- | --- | --- | --- |
|  | Sensitivity | Specificity | Accuracy | F-measure |
| **KNN** | 0.92 | 0.33 | 0.81 | 0.92 |
| **SVM** | 0.97 | 0.6 | 0.9 | 0.97 |
| **NB** | 0.7 | 0.93 | 0.74 | 0.7 |
| **DT** | 0.92 | 0.73 | 0.89 | 0.92 |
| **RF** | 0.97 | 0.53 | 0.89 | 0.97 |

KNN: k-nearest Neighbors; SVM: Support Vector Machine; NB: Naive Bayes; DT: Decision Trees; RF: Random Forest.

**Figure 1.**
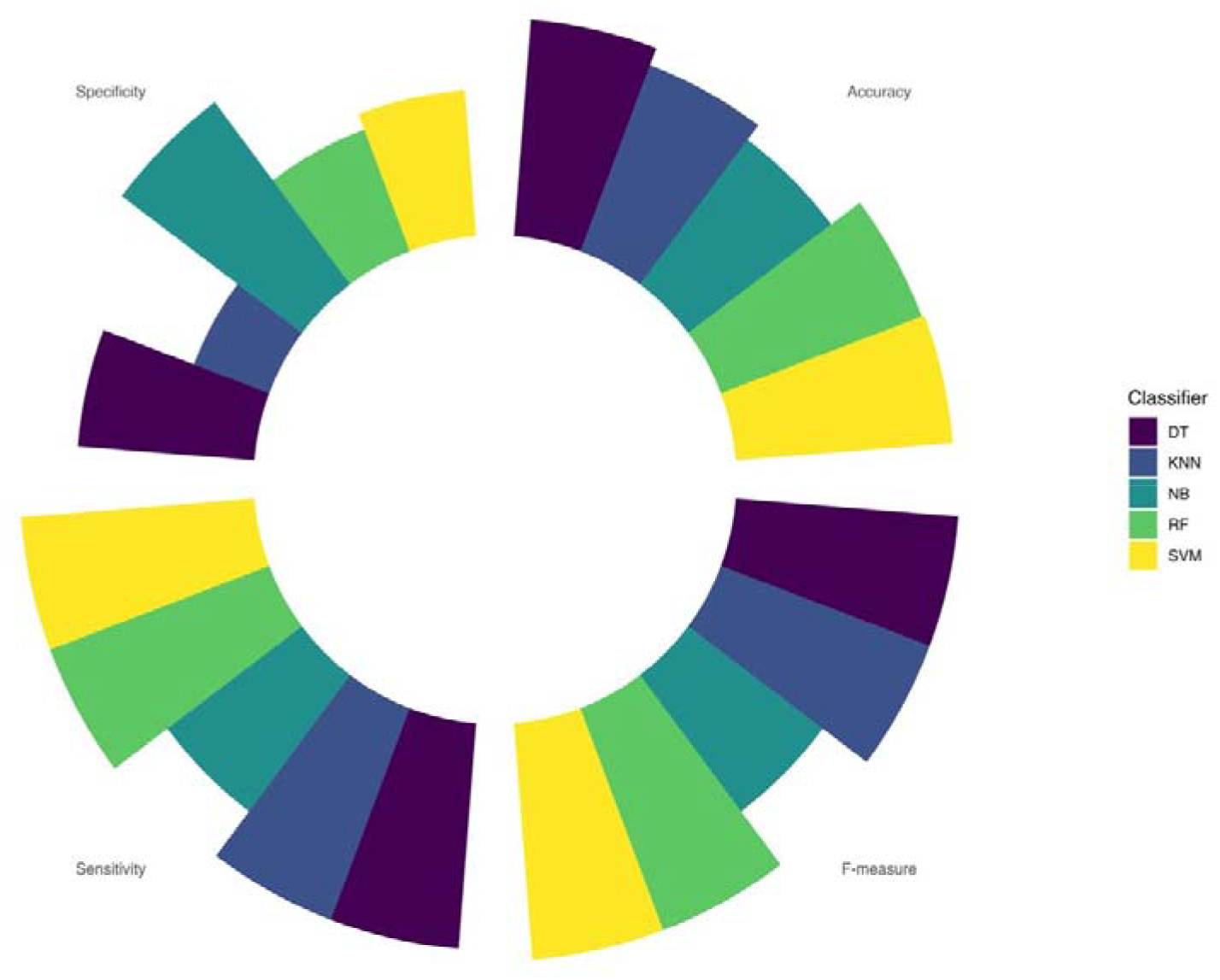
Comparison of F-measure, sensitivity, specificity, and accuracy metrics between KNN, SVM, DT, and RF ML models for predicting outcomes after Alteplase therapy in ischemic stroke patients.

When evaluating the accuracy, SVM outperformed the others with a score of 0.9, suggesting its strong overall predictive capability while NB showed the lowest effectiveness in producing correct overall predictions. The F-measure revealed that both SVM and RF achieved the highest validation score of 0.97. this indicates that these models excel in terms of sensitivity and maintain a favorable balance between precision and recall, making them highly suitable for practical applications.

### 3.2. Feature Selection

To identify the top features that significantly have a central role in predicting the potential effect of Alteplase therapy in ischemic stroke patients, we adopted a backward elimination strategy. We created a modified version of the dataset by omitting each feature and retraining the models with this new dataset. The performance of each retrained model was then assessed using the accuracy metric. Comparing the performance of each modified model with the baseline model, each removed feature was mapped to its corresponding accuracy. Features with a significant drop in a model’s performance due to its elimination were identified as critical while those with minimal impact were eliminated. Figure 2 visually represents the importance of the identified features, ranked from most to least impactful. Features at the uppermost part of the plot signify the greatest role in enhancing predictive accuracy, while those at the bottom are deemed less relevant.

**Figure 2.**
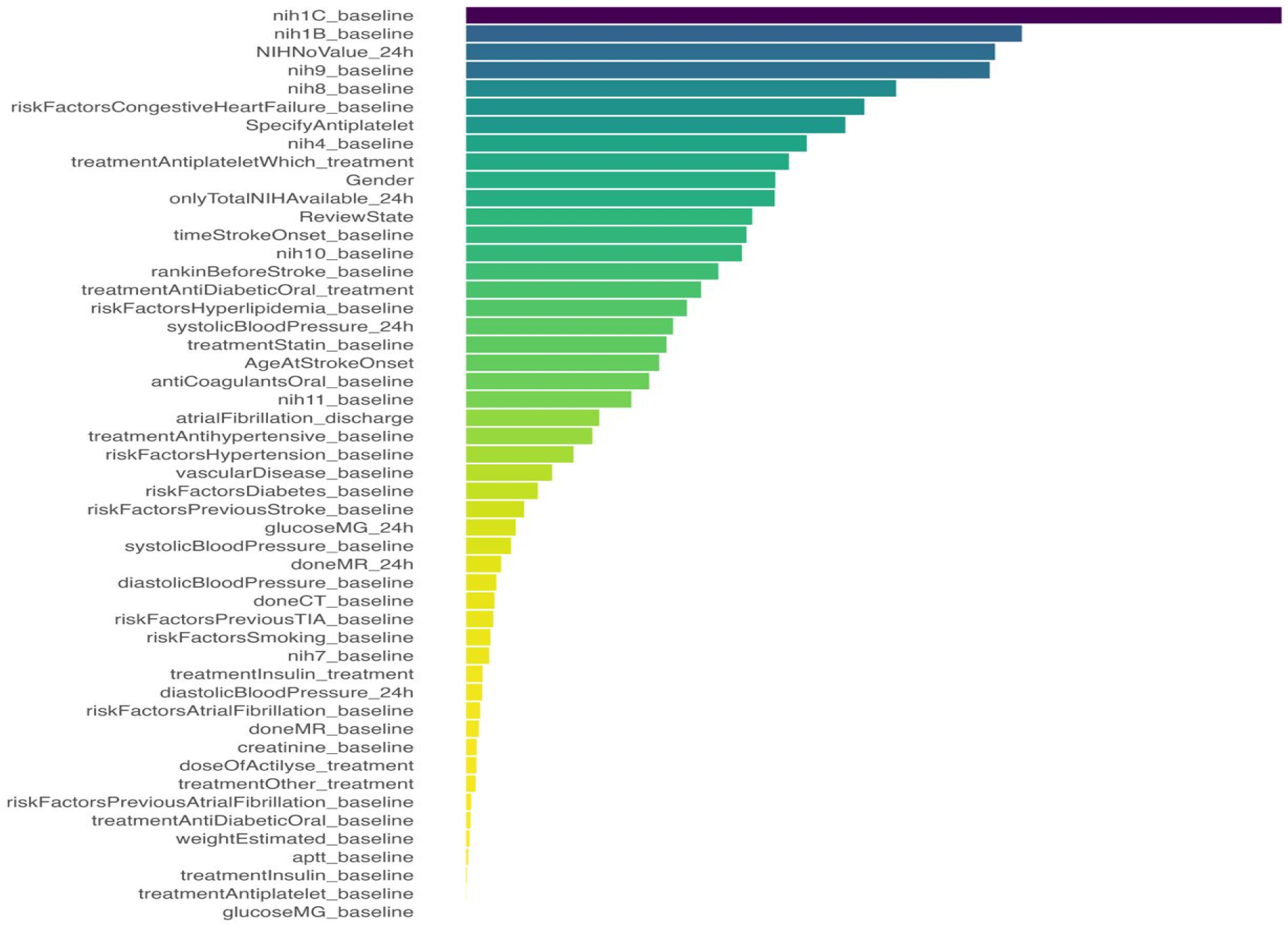
Visualization of key features influencing outcomes after Alteplase therapy in ischemic stroke patients. The color gradient illustrates feature importance, with purple and blue denoting high significance, while orange to yellow indicates lower relevance.

Among the most important features identified, NIH1C (LOC commands (eye and hand movements)), NIH1B (LOC questions (birthday and age recall)), and NIH_noValue (the absence of any stroke characteristics) had the highest scores, indicating their importance in accurately predicting patient outcomes. In addition, NIH9 (Best language (object identification, repetition, and speech production)), NIH8 (Sensory function (sensory loss evaluation)), congestive heart failure, anti-platelet medication, NIH4 (facial palsy), type of antiplatelet treatment, and gender were other significant features. These top ten features were selected for training the final models.

On the other hand, the glucose level and the antiplatelet treatment were found to be the least significant features. The feature selection results underscore the centrality of specific neurological assessment in predicting outcomes, emphasizing the potential for these features to guide clinical decision-making.

### 3.4. Final Model Training

After selecting the top ten features identified as significant predictors, we proceeded to train five ML models including KNN, SVM, NB, DT, and RF, to predict outcomes for ischemic stroke patients receiving Alteplase therapy. Several evaluation metrics including sensitivity, specificity, accuracy, and F1-measure were then calculated.

It is noteworthy that, while a decrease in some measures was observed across all models after feature selection, the majority of models exhibited unchanged or even enhanced values. The detailed performance of each ML model is shown in Figure 3.

**Figure 3.**
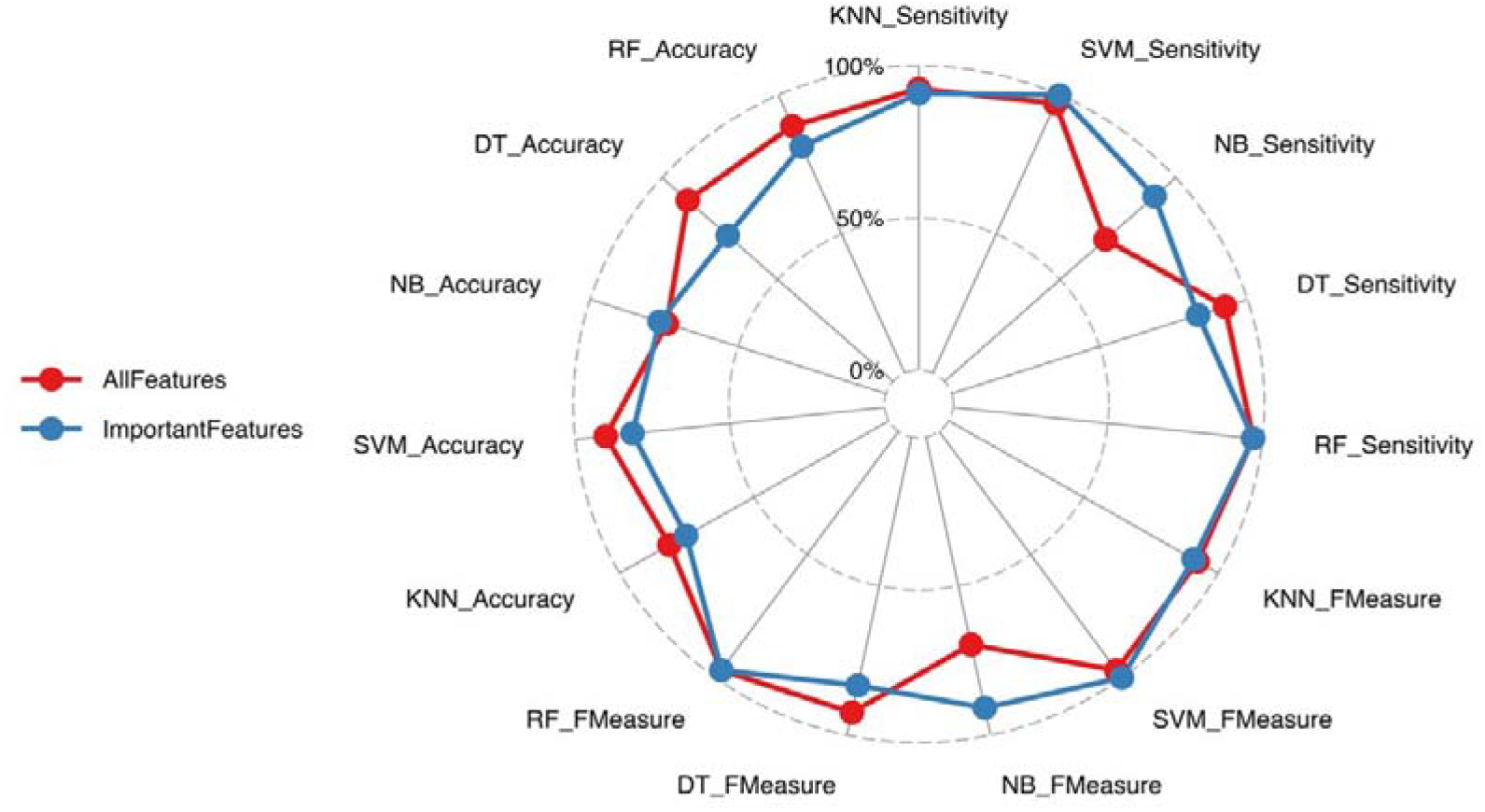
Radar plots depicting the performance of the various algorithms before and after important feature selection for predicting outcomes after Alteplase therapy in ischemic stroke patients.

## 4. Discussion

In this study, we developed and validated machine learning models for predicting the potential effect of Alteplase therapy in ischemic stroke patients. Our initial approach involved constructing models using KNN, SVM, DT, NB, and RF algorithms with fifty variables.

Based on the results, models made by RF and SVM demonstrated acceptable performance compared to other ML algorithms. Accordingly, Kaiting Fan et al. constructed an accurate and personalized secondary prevention ML model, which achieved an area under the curve (AUC) of 0.790 with RF compared to other models including SVM, NB, and logistic regression (LR), particularly in delivering accurate and timely prediction for stroke patients following 30 days intravenous Alteplase treatment [31].

Similarly, Ahmad A. Abujaber et al., found that the SVM model, with an AUC of 0.72, outperformed other ML models in predicting prognostic outcomes for ischemic stroke patients who had received thrombolytic (fibrinolytic) treatment [32]. In a separate study by Zheng Ping et al., RF, LR, and SVM emerged as the top three algorithms among various ML models, demonstrating the highest accuracy for predicting long-term outcomes in stroke patients post-intravenous thrombolysis [33]. Furthermore, Hamed Asadi, et al., reported that the SVM model displayed significant predictive capability for acute ischemic stroke after intra-arterial therapy, achieving a root mean squared error of 2.064 [34]. These findings highlight the potential of ML models to uncover complex relationships and hidden patterns among a wide range of input variables, thereby facilitating the most accurate predictions possible in clinical settings [35].

However, within the medical field, employing a large number of features in predictive modeling may not always be the most effective approach. Gathering extensive data can pose a significant challenge for practitioners, particularly in clinical settings where guidelines may vary and not all information is constantly accessible. Moreover, including numerous features can complicate analyses, leading to increased computational costs, increased complexity, and reduced interpretability of the models [36, 37]. Therefore, simpler models are easier to interpret, more widely applicable, and better suited for practical use. However, it is crucial to ensure that essential variables are not ignored in the pursuit of model simplicity [38].

To balance model parsimony with the inclusion of essential features, we employed a backward elimination approach, a type of wrapper strategy [39], to select the most significant predictors while ensuring that our model remains relatively efficient and interpretable.

Among the most important features identified, NIHSS scores including NIH1C (LOC commands (eye and hand movements)), NIH1B (LOC questions (birthday and age recall)), and NIH_noValue (the absence of any stroke characteristics), NIH9 (Best language (object identification, repetition, and speech production)), and NIH8 (Sensory function (sensory loss evaluation)) had the highest scores, indicating their importance in accurately predicting patient outcomes.

Baseline NIHSS scores, which were identified by the ML algorithms of the present study as important key features, serve as a standardized tool for assessing the potential effects of Alteplase therapy in ischemic stroke patients [40, 41]. Congestive heart failure, anti-platelet medication, NIH4 (facial palsy), type of antiplatelet treatment, and gender were other significant features.

Prior studies further support the relevance of these findings. For example, Mingfeng Zhai et al. identified a correlation between age and baseline NIHSS scores with increased long-term mortality in stroke patients [42]. Similarly, Yinglei Li et al., emphasized that factors such as posterior circulation stroke, elevated NIHSS scores at admission, and specific laboratory parameters (such as homocysteine levels) are significant predictors of post-Alteplase outcomes [43]. Additionally, Yanan Xu et al., found that baseline NIHSS, systolic blood pressure at admission, and the neutrophil-to-lymphocyte ratio (NLR) were critical features for predicting hemorrhage transformation in acute ischemic stroke after alteplase, with the baseline NIHSS score serving as an independent risk factor [44].

Consistent with previous research, a high baseline NIHSS score, which indicates severe or diffuse neuron impairment due to ischemic stroke, was associated with poor outcomes [45]. Gender was also identified as a significant factor influencing treatment outcomes as Kaiting Fan MS found that age along with NIHSS scores at admission are important features for enhancing ML-driven ML predicting [31]. Furthermore, Carolyn Breauna Sanders et al. highlighted heart failure as a critical predictor of unfavorable outcomes in ischemic stroke patients, noting that those with both ischemic stroke and heart failure were associated with improved ambulatory status post-Alteplase therapy [46].

Understanding these critical factors provides valuable insight into identifiable risk factors that can be managed to enhance the care of ischemic stroke patients after Alteplase therapy. Furthermore, the assessment of Alteplase therapy during hospitalization is essential for optimal patient management. Clinicians often focus predominantly on laboratory-based indicators while sometimes underestimating non-laboratory characteristics. Objective assessments of disease severity are vital for medical decision-making. Our findings underscore the importance of evaluating the functional status of ischemic stroke patients following Alteplase therapy, as this can significantly impact clinical outcomes.

## 5. Conclusion

This study underscores the potential of ML models, especially random forest (RF), in predicting outcomes for ischemic stroke patients under Alteplase therapy. Its ability to accurately classify outcomes while maintaining sensible sensitivity and specificity suggests it is a useful tool for clinicians aiming to improve patient care.

Here’s a sample statement for including ethical review information in a manuscript:

## Data Availability

All data produced in the present study are available upon reasonable request to the authors

## 6. Ethics Approval Statement

This study was conducted in compliance with ethical guidelines and was approved by Ethics Committee of Zanjan University of Medical Sciences, Zanjan, Iran. Ethics approval was obtained prior to data collection, with approval number ZUM-4038. All participants provided informed consent, and their confidentiality and rights were protected throughout the study.

